# Automated lung sound analysis using the LungPass platform: A sensitive and specific tool for identifying lower respiratory tract involvement in COVID-19

**DOI:** 10.1101/2021.07.08.21260125

**Authors:** Elena A. Lapteva, Olga N. Kharevich, Victoria V. Khatsko, Natalia A. Voronova, Maksim V. Chamko, Irina V. Bezruchko, Elena I. Katibnikova, Elena I. Loban, Mostafa M. Mouawie, Helena Binetskaya, Sergey Aleshkevich, Aleksey Karankevich, Vitaly Dubinetski, Jørgen Vestbo, Alexander G. Mathioudakis

**Affiliations:** Belarusian State Medical Academy of Postgraduate Education, Minsk, Belarus; 5th City Clinical Hospital, Minsk, Belarus; Minsk Regional Tuberculosis Dispensary, Minsk Region, Belarus; Minsk Regional Pediatric Clinical Hospital, Minsk Region, Belarus; Minsk Clinical Center of Phthisiopulmonology, Minsk, Belarus; Healthy Networks OU, Tallinn, Estonia; Division of Infection, Immunity and Respiratory Medicine, School of Biological Sciences, The University of Manchester, Manchester Academic Health Science Centre, UK; The North West Lung Centre, Wythenshawe Hospital, Manchester University NHS Foundation Trust, Manchester, UK

**Keywords:** COVID-19, Lung sounds, Auscultation, pneumonia

## Abstract

**Background:** Lower respiratory tract (LRT) involvement, observed in about 20% of patients suffering from coronavirus disease 2019 (COVID-19) is associated with a more severe clinical course, adverse outcomes and long-term sequelae. Early identification of LRT involvement could facilitated targeted and timely interventions that could alter the short- and long-term disease outcomes. The LungPass is an automated lung sound analysis platform developed using neural network technology and previously trained. We hypothesised that the LungPass could be used as a screening tool for LRT involvement in patients with COVID-19.

**Methods:** In a prospective observational study involving 282 individuals with presenting in the emergency department with a strong clinical suspicion of COVID-19 and imaging findings consistent with COVID-19 LRT involvement (25.5% had concomitant hypoxia), and 32 healthy controls, we assessed the sensitivity and specificity of the LungPass in identifying LRT involvement in COVID-19. We also compared the auscultatory findings of the LungPass compared to a chest physician using a traditional, high-quality stethoscope.

**Results:** Among individuals with COVID-19 LRT involvement, the LungPass identified crackles in at least one auscultation site in 93.6% and in two or more points in 84%. Moreover, the LungPass identified any abnormal lung sound (crackles or wheeze) in at least one auscultation site in 98.6% and in at least two points in 94% of the participants. The respective percentages for the respiratory physicians were lower.

Considering the presence of any added abnormal sound (crackles or wheeze) in at least two auscultation points as evidence of LRT involvement, LungPass demonstrated a sensitivity of 98.6% (95% confidence intervals [CI]: 96.4%-99.6%) and a specificity of 96.9% (95% CI: 83.8%-99.9%) in identifying COVID-19 LRT involvement.

**Conclusion:** This exploratory study suggests the LungPass is a sensitive and specific platform for identifying LRT involvement due to COVID-19, even before the development of hypoxia.

## Short report

Lower respiratory tract (LRT) involvement, observed in about 20% of patients suffering from coronavirus disease 2019 (COVID-19), is associated with a more severe clinical course, adverse outcomes, and long-term sequelae^1,2^. By pointing out people at risk of deterioration, early identification of LRT involvement could facilitate targeted and timely administration of treatments that could alter the short- and long-term disease outcomes^3^. While imaging represents the gold standard diagnostic test for LRT involvement, it is associated with a potentially avoidable radiation burden and may not be easily accessible in some treatment settings, such as primary care^4^. On the other hand, oxygen desaturation appears to be a specific, but not sensitive marker, since ground glass changes or consolidation are often observed in the absence of hypoxia^5,6,7^. The sensitivity of chest auscultation in identifying LRT involvement has been evaluated in limited populations and varies^8,9^, possibly to some extent due to variable skill among the assessors.

The LungPass, an automated lung sound analysis platform consisting of an electronic wireless stethoscope paired with a mobile phone application, could standardize auscultation process by limiting observer’s bias. The LungPass algorithm was developed using neural network technology and trained using sequential derivation sets of lung sound recordings. Next, the performance properties of the algorithm were evaluated in a separated validation cohort of 200 sound recordings. All the lung sound recordings used were adjudicated by a panel of expert respiratory physicians. Based on the validation cohort, the LungPass can identify normal lung sounds with a sensitivity of 96.9% and a specificity of 90%, crackles (92.5%, 82.5%) and wheeze (99.4%, 90.0%), as well as identifying artifacts and heart sounds (the development process will be reported separately).

We hypothesized that the LungPass could be used as a screening tool for LRT involvement in patients with COVID-19. In a prospective observational study that was conducted in the 5^th^ City Clinical Hospital and the Minsk Regional Tuberculosis Dispensary (Minsk Region, Belarus), we evaluated the sensitivity of lung sounds assessed by a respiratory physician or by the LungPass in identifying LRT involvement in patients with COVID-19. The study included 282 individuals presenting in the emergency department with a strong clinical suspicion of COVID-19 and imaging findings consistent with COVID-19 LRT involvement (ground glass opacities and/or consolidation). A confirmatory PCR result was available in 72.3% of the participants. Sequential chest auscultation in 11 pre-specified points (figure 1) was conducted using the LungPass followed by a chest physician using a high-quality stethoscope. Lung sounds in each auscultation site were recorded as normal breathing sounds, crackles, wheeze, cardiac sounds or artifacts (the last two types of sounds were excluded). To estimate the performance characteristics of the LungPass (sensitivity and specificity), we also used it to auscultate 32 consecutive patients admitted to the hospital with non-respiratory problems.

**Figure 1.**
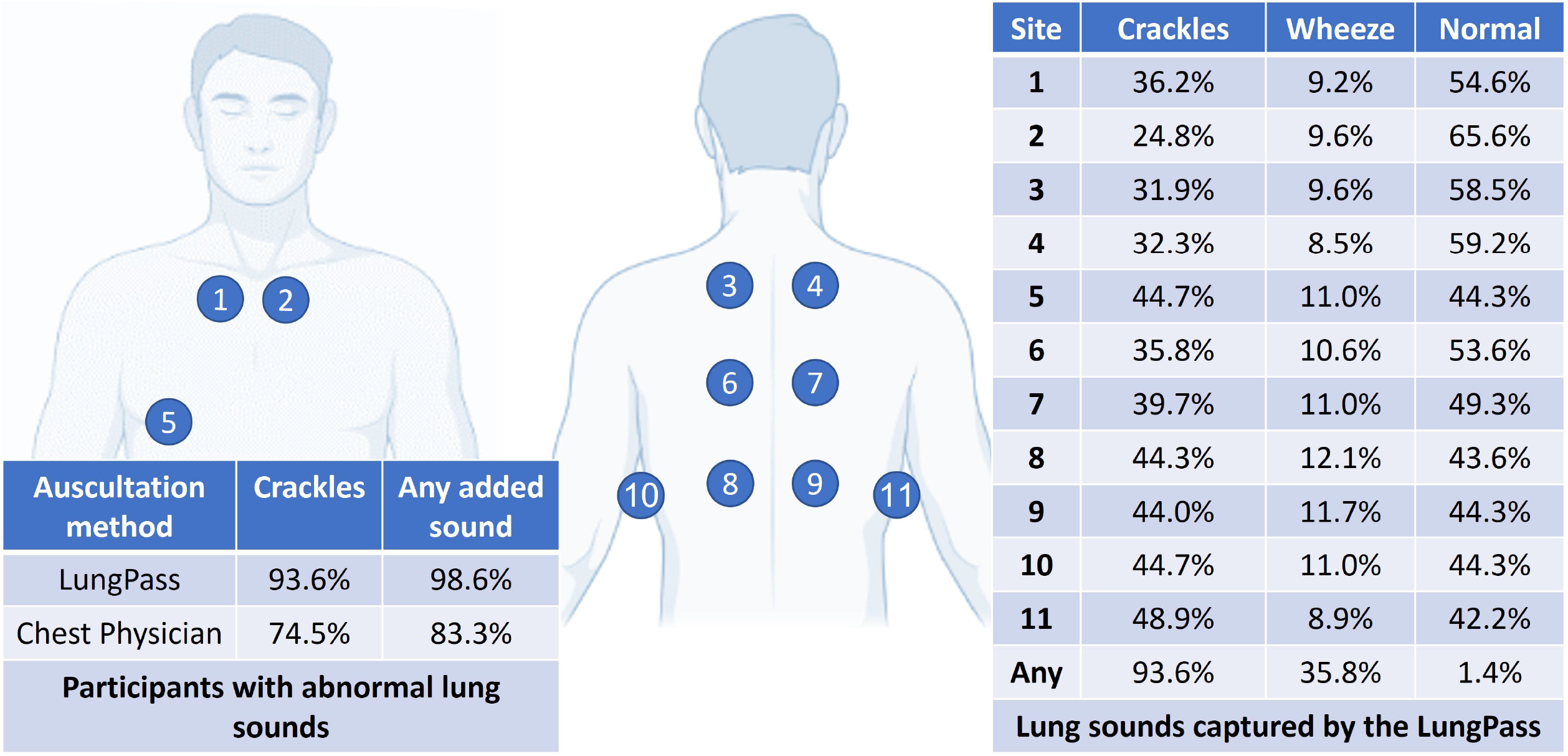
Auscultation points captured by the LungPass and Chest Physicians and the respective findings from participants with COVID-19 and LRT involvement.

The mean age of the participants in the COVID-19 group was 60.2 (standard deviation: 14.5) years and 56% were females. Pre-existing chronic respiratory disorders were reported by 9.2% of these participants (mostly COPD, 6.7%). The median duration of symptoms at recruitment was 6 days (interquartile range: 3-7 days). Most frequent presenting symptoms included fatigue (69.9% of the participants), fever (66.7%), cough (63.8%) and breathlessness (35.8%). Hypoxia (oxygen saturation less than 94%) was observed only in 25.5% of the participants, despite all having radiographic evidence of COVID-19, including ground glass changes (97.9%) and/or consolidation (20.2%).

Overall, crackles were observed more frequently in the lower respiratory fields (figure 1). The LungPass identified crackles in at least one auscultation site in 93.6% and in two or more points in 84.0% of the participants with COVID-19. The respective figures for respiratory physicians were 74.5% and 67.7%. Moreover, the LungPass identified any added abnormal lung sounds (crackles or wheeze) in at least one auscultation site in 98.6% and in at least two points, in 94.0% of the participants, while the respective percentages for the respiratory physician were 83.3% and 79.1%.

Among the 3,102 auscultation sounds (11 points x 282 patients), 533 (17.2%) were deemed abnormal by the LungPass, but normal by the chest physician, while the opposite finding was observed for 175 (5.6%) sounds. Interestingly, abnormal lung sounds were identified in one of the two most proximal, ipsilateral auscultation points in 73.6% and 86.3% of cases in each of the previously described groups, suggesting that the divergence between clinicians and LungPass characterization of the lung sounds is more likely to represent limited sensitivity to added abnormal lung sounds (false negatives), or differences in the positioning of the stethoscope, rather than limited specificity. Within 1,075 lung sounds that were deemed abnormal both by the clinician and LungPass, there was agreement in 97% of cases regarding the type of the abnormality (crackles or wheeze).

The mean age of the control participants was 46.1 (standard deviation: 18.1) years, 34.4% were females and none had underlying chronic respiratory conditions. The LungPass only identified abnormal lung sounds (crackles or wheeze) in one of the control patients, in two auscultation points. Based on our data, if the presence of any added, abnormal sound was considered evidence of LRT involvement, LungPass would have a sensitivity of 98.6% (95% confidence intervals [CI]: 96.4%-99.6%) and a specificity of 96.9% (95% CI: 83.8%-99.9%). However, our previous data has suggested the presence of some false positive lung sound readings. To limit the risk of false positive readings, we could therefore consider the presence of any added abnormal lung sounds in at least two auscultation points as evidence of LRT involvement and in this case the sensitivity of LungPass is estimated to be 94.0% (95% CI: 90.5% - 96.5%) and the specificity 96.9% (95% CI: 83.8%-99.9%).

All results remained unchanged in a sensitivity analyses only including patients with COVID-19 confirmed by a positive PCR.

These findings suggest the LungPass is a sensitive and specific platform for identifying LRT involvement due to COVID-19, even before the development of hypoxia. However, a limitation of this exploratory study was the control group consisting of patients without any acute respiratory problem, rather than patients with confirmed COVID-19 without LRT involvement (with a clear chest CT). Unfortunately, we did not have access to such individuals, since only patients with more severe COVID-19 reached the secondary care setting where this study was conducted during the COVID-19 pandemic. Therefore, future studies are needed to assess the performance characteristics of the LungPass for COVID-19 in a primary care setting.

Several lung sound recording devices have been used to facilitate auscultation during the COVID-19 pandemic without compromising clinician’s personal protective equipment^8,9,10^. Moreover, an extensive observational study evaluating the sensitivity and specificity of a different automated lung sound analysis algorithm is ongoing^11^. However, to our knowledge, this is the first exploratory study demonstrating the sensitivity and specificity of an automated lung sound analysis algorithm (the LungPass platform) in identifying LRT involvement in COVID-19. Upon confirmation of our findings in larger, ongoing studies, the LungPass, an affordable, portable lung sound recording device that could be used by non-experts could facilitate telemonitoring and early identification of patients at risk of deterioration and -possibly-guide timely administration of potentially life-saving treatments.

## Data Availability

Access to de-identified patient level data is available upon request to the authors. Lung sounds cannot be shared as they are protected IP owned by the Healthy Network OU

## Acknowledgement

JV and AGM are supported by the National Institute for Health Research Manchester Biomedical Research Centre (NIHR Manchester BRC).

## Notes

**Conflicts of Interest:** All authors have completed the ICMJE uniform disclosure form summarizing any conflicts of interest they may have. EAL, OK, EIL, MMM, HB, SA, AK and VD are employed by Healthy Networks OU. EAL HB, AK, VD and AGM hold shares in Healthy Networks. VVK, NAV, MVC, IB and EIK have received personal fees from Healthy Network, related to this work. Other potential conflicts of interest outside the submitted work are summarized in the attached CoI declaration forms.

### Competing Interest Statement

All authors have completed the ICMJE uniform disclosure form summarizing any conflicts of interest they may have. EAL, OK, EIL, MMM, HB, SA, AK and VD are employed by Healthy Networks OU. EAL HB, AK, VD and AGM hold shares in Healthy Networks. VVK, NAV, MVC, IB and EIK have received personal fees from Healthy Network, related to this work. Other potential conflicts of interest outside the submitted work are summarized in the attached CoI declaration forms. JV reports personal fees from AstraZeneca, grants and personal fees from Boehringer-Ingelheim, personal fees from Chiesi, GSK and Novartis, outside the submitted work; his son works for Chiesi. AGM reports grants from Boehringer Ingelheim outsde the submitted work.

### Funding Statement

This study was funded and sponsored by Healthy Networks.

### Author Declarations

This study was approved by Belarusian State Medical Academy of Postgraduate Education Ethics Committee (Ethics Committee Reference number: №18/21012020) at its meeting held on 06.02.2020 (Protocol № 1/2020).

